# A novel RT-LAMP workflow for rapid salivary diagnostics of COVID-19 and effects of age, gender and time from symptom onset

**DOI:** 10.1101/2021.06.07.21258288

**Authors:** Gerson Shigeru Kobayashi, Luciano Abreu Brito, Danielle de Paula Moreira, Angela May Suzuki, Gabriella Shih Ping Hsia, Lylyan Fragoso Pimentel, Ana Paula Barreto de Paiva, Carolina Regoli Dias, Naila Cristina Vilaça Lourenço, Beatriz Araujo Oliveira, Erika Regina Manuli, Marcelo Andreetta Corral, Natale Cavaçana, Miguel Mitne Neto, Maria Mirtes Sales, Luiz Phellipe Dell’ Aquila, Alvaro Razuk Filho, Eduardo Fagundes Parrillo, Maria Cássia Mendes-Corrêa, Ester Cerdeira Sabino, Silvia Figueiredo Costa, Fabio Eudes Leal, Germán Gustavo Sgro, Chuck Shaker Farah, Mayana Zatz, Maria Rita Passos-Bueno

**Affiliations:** Centro de Pesquisa sobre o Genoma Humano e Células-Tronco (HUG-CELL), Instituto de Biociências, Universidade de São Paulo (USP) - São Paulo, Brazil; Instituto de Medicina Tropical, Universidade de São Paulo (USP) - São Paulo, Brazil; Grupo Fleury, Research and Development - São Paulo, Brazil; Instituto de Ensino e Pesquisa Prevent Senior - São Paulo, Brazil; Universidade Municipal de São Caetano do Sul (USCS) - São Paulo, Brazil; Instituto de Química, Universidade de São Paulo (USP) - São Paulo, Brazil

**Keywords:** LAMP assay, 2019 Novel Coronavirus, saliva, molecular diagnostics

## Abstract

**Objectives:** Rapid diagnostics is pivotal to curb SARS-CoV-2 transmission, and saliva has emerged as a practical alternative to naso/oropharyngeal (NOP) specimens. We aimed to develop a direct RT-LAMP workflow for viral detection in saliva, and to provide more information regarding its potential in COVID-19 diagnostics.

**Methods:** Clinical and contrived specimens were used to screen/optimize formulations and sample processing protocols. Salivary viral load was determined in symptomatic patients to evaluate clinical performance (n = 90) and to characterize saliva based on age, gender and time from onset of symptoms (n = 49).

**Results:** The devised workflow achieved 93.2% sensitivity, 97% specificity, and 0.895 Kappa for salivas containing >10^2^ copies/μL. Further analyses in saliva showed peak viral load in the first days of symptoms and lower viral loads in females, particularly among young individuals (<38 years). NOP RT-PCR data did not yield relevant associations.

**Conclusions:** This novel saliva RT-LAMP workflow can be applied to point-of-care testing. This work reinforces that saliva better correlates with transmission dynamics than NOP specimens, and reveals gender differences that may reflect higher transmission by males. To maximize detection, testing should be done immediately after symptom onset, especially in females.

**HIGHLIGHTS:**

- Development of DGS, a dithiothreitol/guanidine-based solution for stabilization of the viral genome that increases sensitivity for SARS-CoV-2 detection in saliva;
- Rapid, cost-effective RT-LAMP assay workflow for viral detection in saliva without need of RNA extraction;
- Insights into the differences in viral load between saliva and naso-oropharyngeal specimens, and correlation with age, gender and time from symptom onset;

## INTRODUCTION

Molecular diagnostics of the novel coronavirus (SARS-CoV-2) amidst the COVID-19 pandemic has been crucial to monitor infection dynamics and prevent spread of the disease. The gold standard has been Real-Time Polymerase Chain Reaction (RT-PCR), which is performed on nasopharyngeal and oropharyngeal (NOP) specimens that pose discomfort to patients and require specialized materials and trained healthcare professionals for collection. In addition, RT-PCR typically requires viral inactivation followed by a lengthy RNA extraction/isolation step, further complicating diagnostic workflows and increasing turnaround time for reporting results. Faster and simplified protocols for viral detection are desirable to curb transmission, especially in point-of-care settings and places that lack infrastructure, access to material, or financial means. Reverse transcription loop-mediated isothermal amplification (RT-LAMP) has emerged as a viable, affordable alternative to RT-PCR, since it allows rapid and direct detection of pathogens without nucleic acid extraction and sophisticated equipment (1).

Although SARS-CoV-2 is transmitted via infected saliva droplets and aerosols, attention has been focused on the upper airway tract rather than the oral cavity. Recently, viral shedding has been observed in the salivary glands and oral mucosa, further implicating saliva in infection and transmission (2). Saliva has been considered a suitable alternative specimen for COVID-19 molecular diagnostics and to monitor viral spread, as it is easily accessible and can be collected by unsupervised patients into simple airtight vessels, diminishing costs and risk of transmission (3). Implementation of salivary diagnostics must account for some issues, such as poor understanding of the viral biology in the oral cavity and viral load dynamics across individuals, which is important to determine the optimal test window to detect SARS-CoV-2. Furthermore, development of robust protocols for viral detection in saliva may be useful for diagnosis of other respiratory pathogens.

Here, we describe a novel workflow for RT-LAMP-based detection of SARS-CoV-2 that includes an RNA stabilization solution to prepare saliva specimens without RNA isolation. This workflow stabilizes viral RNA, allows sample manipulation without biosafety rooms and cabins, and shows 93.2% sensitivity for viral loads above 10^2^ μL of saliva, 97% specificity, and 0.895 Kappa coefficient. We also provide insights into viral load differences between saliva and NOP swab specimens and relate them to gender, age, and time from onset of symptoms, further elucidating the diagnostic performance of saliva and bringing forth recommendations to maximize chances of detection. This rapid and efficient workflow is suitable for COVID-19 diagnostics in both centralized or point-of-care settings, and may be of particular value for use in places lacking sophisticated infrastructure.

## METHODS

### Subjects

This project was approved by the Ethics Committee of Instituto de Biociências, Universidade de São Paulo, Brazil (accession number 31655320.0.0000.5464), and involved the collaboration with several groups in order to gain access to anonymized clinical samples from individuals with respiratory symptoms:

a. Crude saliva samples from 26 symptomatic individuals were collected in August 2020 by two clinical laboratories (Instituto de Ensino e Pesquisa Prevent Senior and Grupo Fleury) and one research group (Instituto de Medicina Tropical, Universidade de São Paulo - IMT-USP), and sent to our laboratory in ice on the same day. These individuals have tested positive for SARS-CoV-2 in RT-PCR of nasopharyngeal/oropharyngeal (hereafter referred as NOP) specimens by those institutions, and Ct values were shared whenever necessary.
b. Crude saliva and NOP samples from 131 symptomatic individuals were collected in January-February 2021 at Universidade Municipal de São Caetano do Sul/IMT-USP (4) (51 individuals for characterization of the diagnostic yield of saliva compared to NOP swabs - 29 females and 22 males - and 80 for the blind study - 48 females and 32 males). Saliva samples were aliquoted locally and transported to our laboratory in dry ice, while NOP specimens were tested for SARS-CoV-2 by RT-PCR in each institution, which shared Ct results and clinical data whenever necessary.

### Saliva collection

Briefly, individuals were asked not to eat for at least 30 minutes before collection of 3 mL of saliva in sterile, nuclease-free 15 mL conical tubes. Immediately post-collection, saliva was heat-inactivated by incubation at 95°C.

### DGS preparation

We developed a solution (detailed information is described in the ‘Results’ section), named as ‘DGS’ (DTT/GuHCl solution), that prevents degradation of viral RNA and provides a better stabilization of the viral genome for RT-LAMP reactions. DGS contains 30 mM Tris-HCl pH 8.0, 600 mM GuHCl and 200 mM DTT, diluted in nuclease-free _dd_H_2_O. Inactivated saliva samples are mixed with this solution and incubated at 55°C for 5 minutes prior to RT-LAMP reactions.

### RT-LAMP and rtRT-LAMP reactions

RT-LAMP reactions (12.5 μL total volume) contained 1x WarmStart^®^ Colorimetric LAMP Master Mix (New England Biolabs, #M1800L), a primer set composed of 1.6 μM FIP/BIP internal primers, 0.4 μM LF/LB loop primers and 0.2 μM F3/B3 external primers, and 1.25 μL of DGS:saliva mixture in 1:1 ratio, previously centrifuged at 1,000 x g for 30 seconds). Previously published primer sets targeting different regions of SARS-CoV-2 genome and the human gene *ACTB* were used (Table S1; Broughton et al., 2020; Rabe et al., 2020; Zhang et al., 2020). Reactions were carried out at 65°C for 30 to 40 minutes

For real time analysis of RT-LAMP (rtRT-LAMP), the above reaction was supplemented with 1 μM SYTO^®^-9 DNA binding dye (Thermo Fisher, #S34854) and 0.125 μL Low ROX reference dye (New England Biolabs, #E7638A), and incubated in a QuantStudio 5 qPCR machine (Thermofisher) at 65°C for 30 or 40 minutes (fluorescence signal acquisition at 15-second intervals), followed by a dissociation curve stage from 95°C to 60°C with temperature change rate of 0.1°C/second.

To improve color discrimination, the reaction protocols above was further adjusted to contain 1.32x WarmStart^®^ Colorimetric LAMP Master Mix and 1 μL of DGS:saliva mixture in 2:1 ratio. Time to threshold (Tt) of positivity was defined at threshold = 0.8 relative fluorescence units. Each batch of reactions included positive controls with 1000 and 500 copies of SARS-CoV-2 RNA per reaction. Analysis of amplification plots and melting temperatures (Tm) were used to discriminate non-specific from specific amplifications (specific Tm = median Tm_positive control_ ± 1°C).

### RNA isolation and RT-PCR reactions on saliva samples

SARS-CoV-2 RNA was isolated from crude saliva samples using QIAamp Viral RNA Mini Kit (QIAGEN, #52906), following manufacturer’s recommendations. RT-PCR was performed based on CDC’s protocol, which targets SARS-CoV-2 gene N (Centre for Disease Control and Prevention, 2020). RT-PCR reactions (12 μL total volume) contained TaqMan^®^ Fast Virus 1-Step Master Mix (Thermo Fisher, #4444432) and a primer/probe mix (500 nM forward primer, 500 nM reverse primer, 125 nM probe; 2019-nCoV_N1, IDT #10006600), and were cycled in a QuantStudio 5 qPCR machine as per manufacturer’s recommendations (Thermo Fisher). Absolute quantification of viral RNA copies was performed via standard curve assays with 2019-nCoV_N_Positive Control (IDT, #10006625), in triplicates.

### Simulation of saliva positive controls with SARS-CoV-2 RNA

SARS-CoV-2 RNA was isolated from cell culture pellets (kindly provided by collaborators from Instituto de Ciências Biomédicas, USP), and viral titer was determined via absolute quantification with RT-PCR, as described above. Quantified SARS-CoV-2 RNA was spiked into saliva from a healthy donor (previously processed with 2:1 ratio of DGS:saliva and tested negative for SARS-CoV-2) to produce simulated specimens, and then stored in 10-µL aliquots at -80°C for further use in RT-LAMP reactions.

## RESULTS

### Stabilization of SARS-CoV-2 RNA in saliva

Searching for a solution capable of stabilizing SARS-CoV-2 RNA in saliva, we initially screened 6 formulations containing Proteinase K (namely PK1-6). AVL, a commercial guanidine-based buffer recommended for SARS-CoV-2 inactivation (5), was used as the experimental control, without any heat treatment. Since unprotected viral RNA is rapidly degraded in crude saliva, we simulated samples by first mixing saliva from a healthy donor with the solutions (1:1, v/v) and heating at 65°C for 15 minutes followed by a step at 95°C for 2 minutes, before spiking in SARS-CoV-2 RNA. After sample processing, RNA was isolated and RT-PCR targeting the N gene was performed. We observed that PK6 led to similar Ct values to AVL (23.9 and 23.15, respectively), while no amplification was detected for the remaining PK formulations. Notably, PK6 and AVL were the only solutions that contained guanidine hydrochloride (GuHCl), suggesting that GuHCl is important for stabilizing the viral RNA. PK6 was composed of 800 mM GuHCl, 400 μg/mL PK, 10% Tween 20 (T20) and 30 mM Tris-HCl pH 8.0 (Table 1).

**Table 1:** Solutions used in the initial screening and Ct values after processing.

|  | Composition | Ct |
| --- | --- | --- |
| **AVL** | Guanidinium thiocyanate (50-70%) | 23.15 |
| **PK1** | PK (2 mg/mL) | ND |
| **PK2** | PK (0.4 mg/mL) | ND |
| **PK3** | PK (0.4 mg/mL) + T20 (10%) | ND |
| **PK4** | PK (0.4 mg/mL) + T20 (10%) + Tris-HCl pH 8.0 (30 mM) | ND |
| **PK5** | PK (0.4 mg/mL) + Tris-HCl pH 8.0 (30 mM) | ND |
| **PK6** | PK (0.4 mg/mL) + T20 (10%) + GuHCl (800 mM) + Tris-HCl pH 8.0 (30 mM) | 23.9 |

PK: Proteinase K; T20: Tween 20; GuHCl: guanidine hydrochloride; ND: not detected;

Aiming towards a minimal solution that enables RNA stabilization, several modifications were made to the PK6 formula. The removal of T20 and PK increased the Ct value in relation to AVL (ΔCt = 1.433), while no amplification was detected when substituting GuHCl with PK at a higher concentration (4 mg/mL), RNAse OUT (Thermo Fisher, #10777019) (6) or varying amounts of DTT (see below), further indicating that GuHCl is necessary to stabilize SARS-CoV-2 RNA in saliva (Fig. 1A). By varying the amount of GuHCl in PK6 (1600 mM, 800 mM, 400 mM, 200 mM), we observed that viral RNA could be detected only when employing concentrations above 400 mM, and that 800 mM was sufficient for RNA stabilization (ΔCt ≤ 0.104) in the presence of T20 and PK (Fig. 1B). PK and T20 were then replaced with varying amounts of DTT (50 mM to 200 mM) in combination with lower concentrations of GuHCl (400 mM and 600 mM). While 400 mM GuHCl combined with any amount of DTT led to significantly greater Ct values (ΔCt ≥ 2.572), combining 600 mM GuHCl and 200 mM DTT resulted in a negligible Ct increment in comparison to AVL (ΔCt = 0.148) (Fig. 1C). Furthermore, saliva samples processed with this formulation presented the smallest Ct rise after storage at 8°C for 24 hours (ΔCt = 0.428) (Fig. 1D). Together, these results indicate that heating saliva in this solution (30 mM Tris-HCl pH 8.0, 600 mM GuHCl, 200 mM DTT) at 65°C/15 minutes plus 95°C/2 minutes protects viral RNA from degradation. This DTT/GuHCl solution is hereafter referred to as ‘DGS’.

**Figure 1.**
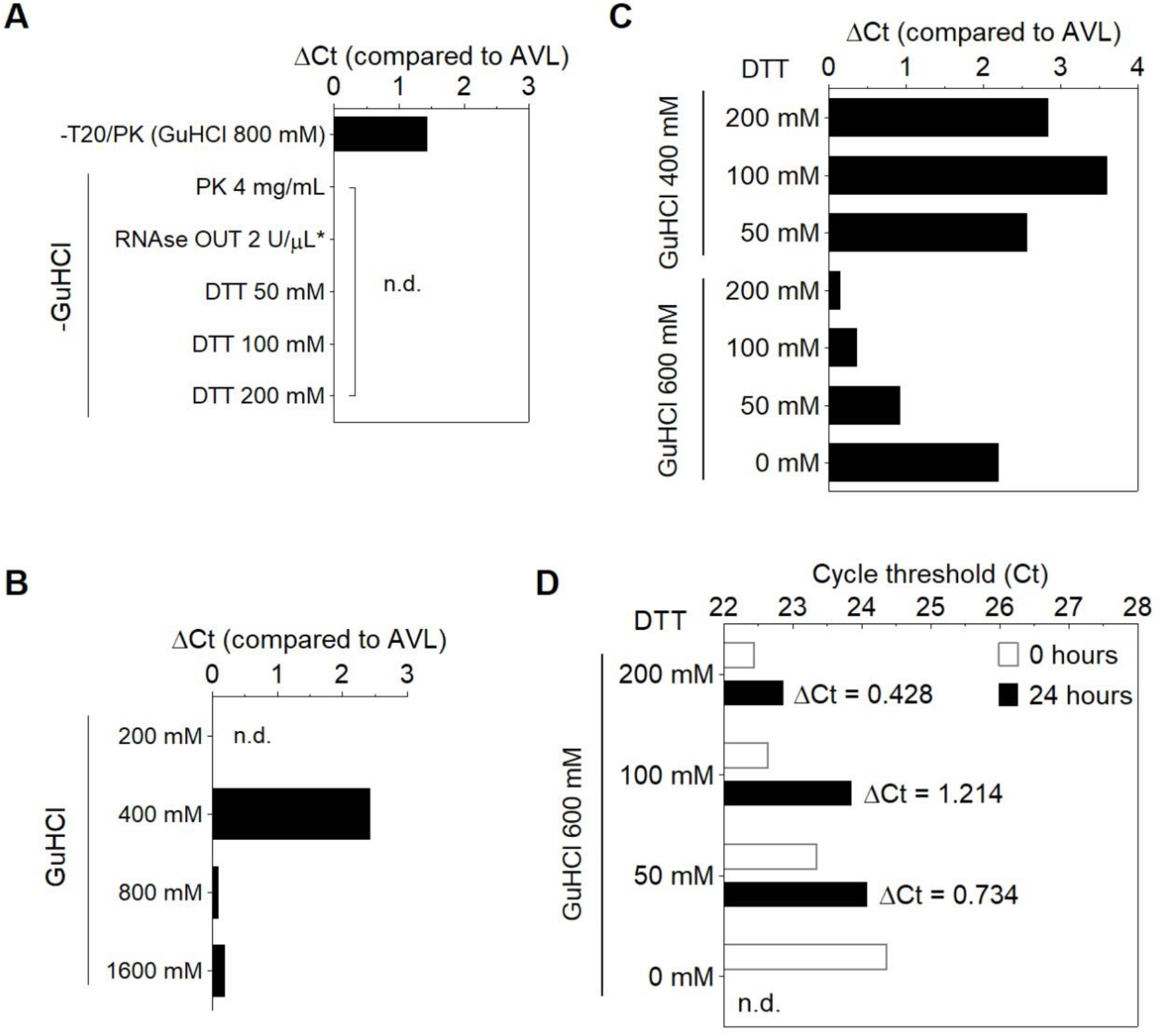
Modifications to the PK6 formula. **A)** Removal of PK and T20 increases Ct value, while substituting GuHCl with PK (4mg/mL), RNAse OUT or DTT results in no amplification. **B)** 800 mM GuHCl is sufficient to stabilize RNA in the presence of PK and T20. (*) The solution containing RNAse OUT was buffered by Tris-EDTA pH 8.0 instead of Tris-HCl (6). **C, D)** Optimization of the stabilization solution. **C)** RNA stabilization is achieved by replacing PK and T20 with 200 mM DTT and reducing GuHCl to 600 mM, and **(D)** this is maintained after storage of processed specimens for 24 hours at 8°C. n.d. = not detected.

### Compatibility of DGS with direct RT-LAMP in saliva

Next, we examined whether DGS is compatible with direct RT-LAMP reactions. For this, we used saliva specimens from 3 individuals confirmed positive for COVID-19 via NOP swab RT-PCR by an external laboratory. In this test, specimens were heated with DGS under a slightly different protocol (55°C/15min followed by 95°C/2min) modified from previous work (7). RT-LAMP was performed with the previously reported primer set N (Table S1) (8) and 10% volume of DGS:saliva (1:1) mix (total reaction volume = 12.5 μL), resulting in a 20-fold dilution of the DGS formulation (Table 2). These 3 samples amplified specifically and changed color from pink to yellow within 30-40 minutes of reaction at 65°C, while the no-template controls (NTCs) remained pink (Fig. 2A), showing that DGS is compatible with colorimetric readouts.

**Table 2:** DGS constituents and carryover into RT-LAMP

|  | **DGS** | **DGS:saliva (1:1)** | **Carryover into RT-LAMP**<br>**(1.25 μL input)** |
| --- | --- | --- | --- |
| **GuHCl** | 600 mM | 300 mM | 30 mM |
| **DTT** | 200 mM | 100 mM | 10 mM |
| **Tris-HCl pH 8.0** | 30 mM | 15 mM | 1.5 mM |
| **Saliva** | - | 100 μL | 0.625 μL |
| **Total volume** | 100 μL | 200 μL | 12.5 μL |

**Figure 2:**
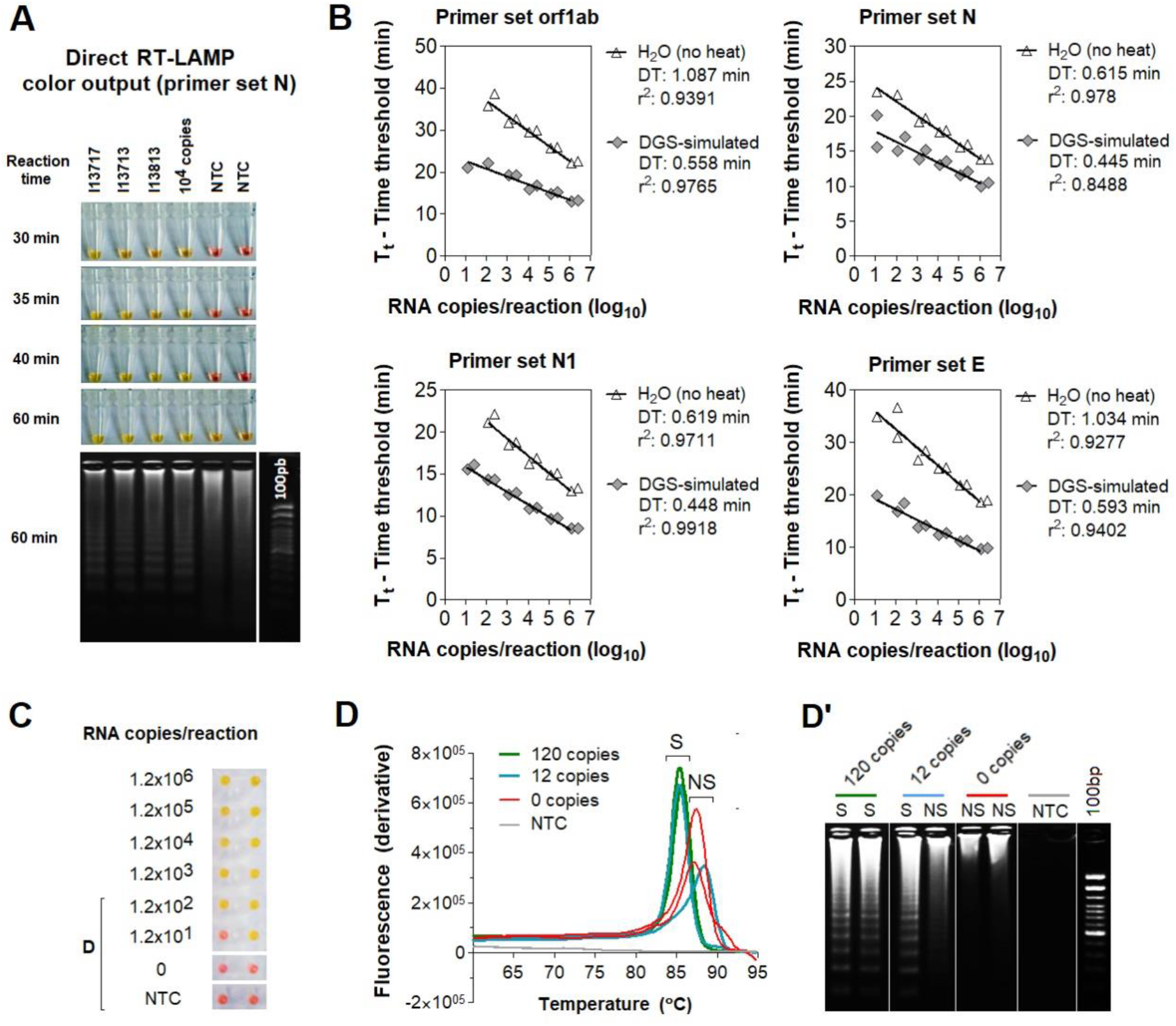
Effect of DGS heating on direct RT-LAMP reactions. **A) Compatibility between direct RT-LAMP and DGS-processed saliva.** After 40-minute incubation at 65°C, colors were visibly distinguishable between the pink NTC and the yellow positive samples. Agarose gel electrophoresis confirmed specific amplification, as band patterns of individuals 13717, 13713 and 13813) matched those of the positive control (10^4^ RNA copies). Nonspecific amplification was observed in NTC after longer incubation (up to 60 minutes). **B-F)** Compatibility with direct rtRT-LAMP. **B)** SARS-CoV-2 RNA serially spiked in DGS-simulated saliva or in H_2_O was used to assemble standard curves for primer sets orf1ab, N, N1 and E, in duplicate reactions. Nonspecific and failed amplifications were observed at 0-120 copies/reaction in some reactions (not shown). Doubling time (DT) values were calculated to assess amplification speed/efficiency (11) for each primer set. Determination coefficients (r^2^) point to high linearity (>0.939) between RNA input and Tt in DGS-simulated saliva, except for primer set N (r^2^ = 0.8488). **C)** Representative color output after rtRT-LAMP (primer set E is shown). **D)** Representative dissociation analysis and gel electrophoresis showing non-specific LAMP products at 0 and 12 copies/reaction (primer set E is shown). NTC (no-template control) reactions were performed with H_2_O.

To characterize the effects of the DGS heat treatment on RT-LAMP, we performed direct real-time RT-LAMP (rtRT-LAMP) with additional 3 published primer sets targeting different regions of the SARS-CoV-2 genome (orf1ab, N1, E ;Table S1) (8–10). Standard curves (1.2 ×10^1^ to 1.2×10^6^ copies/reaction) were made with serially diluted viral RNA spiked into DGS-processed saliva or H_2_O. In DGS-processed samples, rapid and specific amplification was observed for all RNA dilutions. Compared to RNA in H_2_O, DGS-simulated saliva showed higher reaction speed and amplification efficiency for all primer sets, revealed by reduction in time to threshold (Tt) and doubling time (DT) values, respectively (Fig. 2B). In rtRT-LAMP, DGS processing also enabled formation of specific LAMP products and color change to yellow, while nonspecific and failed amplifications remained pink (Fig. 2C). Dissociation analysis and agarose gel electrophoresis showed that specific amplifications produced a single melting peak clearly distinguishable from nonspecific amplifications (Fig. D, D’). Altogether, these results show that sample processing with DGS improves direct RT-LAMP reactions and allows SARS-CoV-2 detection either via endpoint analysis of color output or analysis of amplification and melting curves in rtRT-LAMP.

### Optimization of the DGS workflow

The aforementioned protocols require mixing infected patient samples with DGS before viral inactivation, demanding high biosafety requirements. To overcome this issue, we sought to optimize sample processing to include a heat inactivation step before collection tubes were uncapped and further manipulated. As the starting point for comparison, we tested 3 saliva specimens heat-inactivated at 95°C/20 minutes (protocol A) in the absence of DGS (no-DGS control), resulting in failed amplifications across reaction replicates in rtRT-LAMP. In contrast, subjecting these heat-inactivated specimens to a second heating step with DGS at 55°C/15 minutes (protocol B) improves detection, leading to specific amplification in all replicates. This was also observed when specimens were mixed with DGS and single-heated at 95°C/20 minutes (protocol C) or under the previous protocol (55°C/15 minutes; 95°C/2 minutes; protocol D), suggesting that 2-step heating (protocol B) does not overly affect stabilization of SARS-CoV-2 RNA in saliva (Fig. 3A).

**Figure 3:**
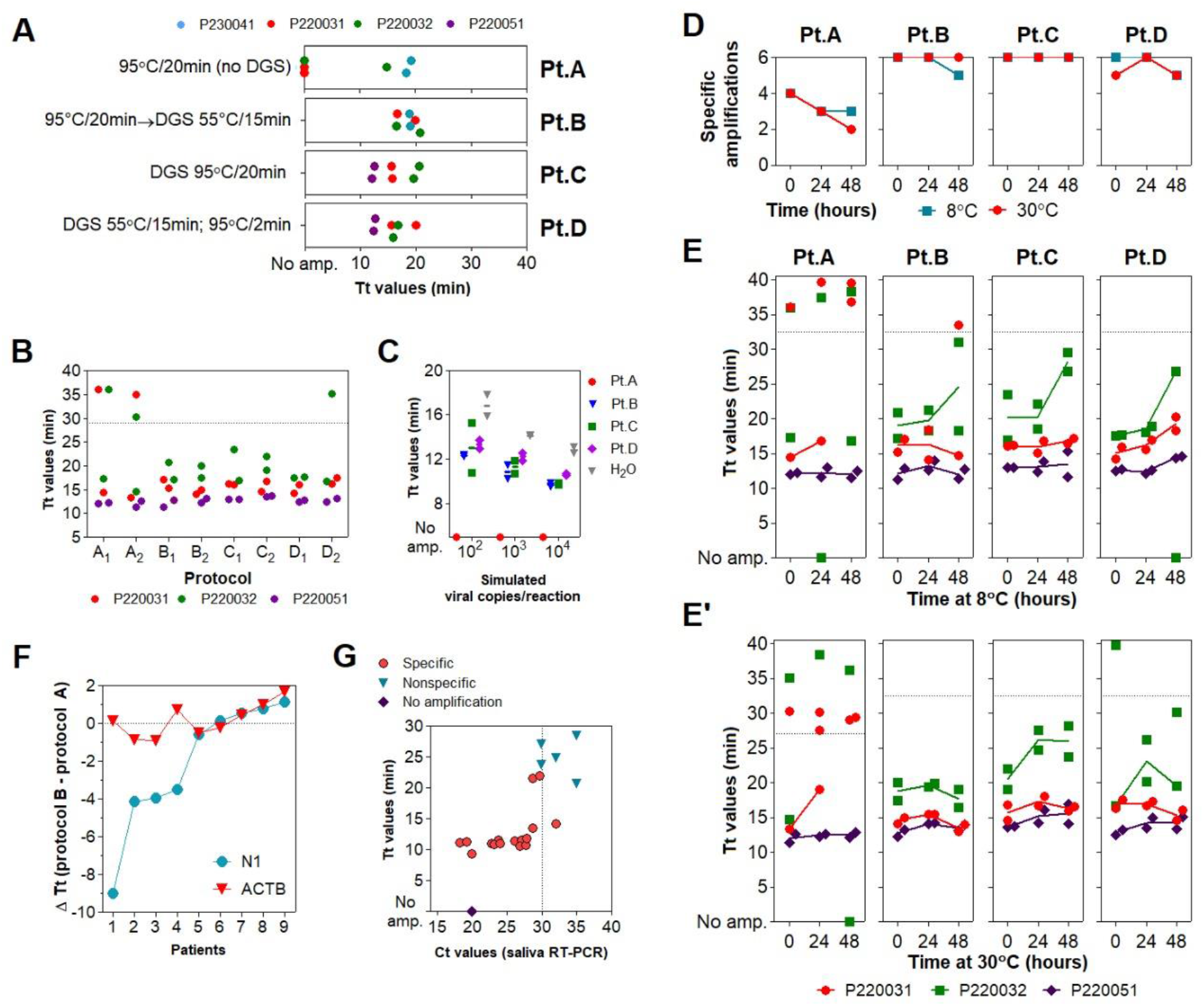
Assessment of different DGS heating protocols via rtRT-LAMP. **A)** rtRT-LAMP results for clinical specimens heat-inactivated (no-DGS control) or processed under 4 DGS protocols (Pt.A-D). Reactions were performed in different days. **B)** rtRT-LAMP results for clinical specimens after protocols A-D, in two parallel experiments. Data points above the dashed line are nonspecific amplifications. rtRT-LAMP was performed in a single reaction plate. **C)** Reaction output of simulated samples spiked with SARS-CoV-2 RNA (10^2^ to 10^4^ copies/reaction). RNAs spiked in H_2_O were used as positive controls. Lines indicate mean Tt values. **D)** Detection rate of the 3 DGS-processed clinical specimens incubated for 48 hours at 8°C and 30°C (n = 6). **E, E’)** rtRT-LAMP results after storage at 8°C **(E)** and 30°C **(E’)**. Data points above the dashed line are nonspecific amplifications. Results shown in D/E/E’ were plotted with data from B (t = 0 hours). **F-G)** Assessment of protocol B in 11 additional specimens via rtRT-LAMP for primer sets N1 and *ACTB*. **F)** Changes in reaction speed of protocol B compared to A were plotted as ΔTt values; negative values indicate gain in reaction speed. Nonspecific and failed amplifications were not included. **G)** Saliva RT-PCR Ct values were plotted against Tt values used in F. Experiments in A-E were performed with n = 3 biological samples (P220031, P220032, and P220041 or P220051). All rtRT-LAMP reactions were carried out in duplicates. Since GuHCl was recently shown to improve speed and sensitivity of RT-LAMP (10), all rtRT-LAMP performed on no-DGS controls (protocol A) were supplemented with 40 mM GuHCl to allow more precise evaluation of the DGS protocols.

We further characterized DGS protocols B, C and D in comparison to the no-DGS control (protocol A). Three saliva specimens were processed under these protocols in 2 parallel experiments. rtRT-LAMP showed no differences in Tt values across protocols B, C, and D. However, we observed less specific amplification events under protocol A (4 out of 6 replicates), while protocols B, C and D achieved specific detection in all replicates, except for one replicate under protocol D (Fig. 3B). Similarly, DGS protocols B, C and D enabled detection in simulated specimens (10^4^, 10^3^ and 10^2^ copies/reaction) without differences in average Tt values, while no amplification was detected under protocol A, indicating that single heat inactivation in the absence of DGS is insufficient to counteract salivary ribonuclease activity (Fig. 3C). To further compare RNA stabilization across protocols, the processed specimens (Fig. 3B) were then incubated at 8°C or 30°C for up to 48 hours. After 24 and 48 hours of incubation at either temperature, the number of specific amplifications under protocol A further decreased, while greater detection rates (≥ 5 out of 6 replicates) were maintained with DGS protocols B, C and D (Fig. 3D). Moreover, these 3 DGS protocols showed no appreciable shifts in average Tt values after 24 hours at 8°C (Fig. 3E), while at 30°C, this was only observed for protocol B (Fig. 3E’). Together, these results suggest that the 2-step protocol B is suitable to process clinical specimens, as it improves detection of SARS-CoV-2 compared to the no-DGS controls, and shows the most robust RNA stabilization effects.

To further assess its performance in rtRT-LAMP, protocol B was applied to 11 additional COVID-19 clinical samples and compared to single heat inactivation without DGS (protocol A). rtRT-LAMP was performed in duplicates with primer set N1 and the human-specific primer set *ACTB* (Table S1) (10). In both methods, specific amplifications for *ACTB* were observed throughout all replicates, while specific amplifications for N1 were observed in at least 1 out of 2 replicates in 9 samples. Differences in reaction speed (ΔTt) were calculated as average *Tt*_*Protocol B*_ *-Tt*_*Protocol A*_. In 4 of these 9 samples, an important reduction in Tt values was observed for protocol B, varying from 3.5 to 9 minutes, while *ACTB* showed no significant Tt fluctuations (Fig. 3F). This indicates that protocol B improves detection rates (Fig. 3A) and reaction speed (Fig. 3F) in clinical saliva, and suggests that the latter effect may be specific to SARS-CoV-2 detection, as *ACTB* showed low ΔTt variability. Finally, viral RNA was extracted from these 11 specimens and RT-PCR Ct values were used to estimate viral load. We observed that SARS-CoV-2 RNA was detected in all specimens with Ct < 30 (8/8), suggesting robustness for high viral loads (Fig. 3G).

### Optimization of color discrimination and sample processing time

We observed that up to 15% specimens showed discordant color output and amplification results after RT-LAMP followed by agarose gel electrophoresis (data not shown). This could be explained by pH variation of saliva or excess of salivary inhibitors impairing amplification efficiency. To ensure appropriate color-based analysis, volumetric adjustments were implemented in the DGS and RT-LAMP protocols. The DGS:saliva ratio was increased to 2:1 to enhance inactivation of salivary nucleases by DTT/GuHCl, while sample input into RT-LAMP reactions decreased from 1.25 µL to 1 µL and the volume of Colorimetric Master Mix increased from 6.25 µL to 8.25 µL (Table 3). These modifications resulted in better color discrimination after RT-LAMP, as color change and viral RNA amplification showed no discrepancies (Fig. 4A). These adjustments were associated with a >95% limit of detection at 750 viral copies/µL in simulated specimens (Fig. 4B).

**Table 3:** DGS constituents and carryover into RT-LAMP after volumetric adjustments.

|  | **DGS** | **DGS:saliva (2:1)** | **Carryover into RT-LAMP**<br>**(1 μL input)** |
| --- | --- | --- | --- |
| **GuHCl** | 600 mM | 400 mM | 32 mM |
| **DTT** | 200 mM | 133.3 mM | 10.66 mM |
| **Tris-HCl pH 8.0** | 30 mM | 20 mM | 1.6 mM |
| **Saliva** | - | 100 μL | 0.33 μL |
| **Total volume** | 200 μL | 300 μL | 12.5 μL |

**Figure 4:**
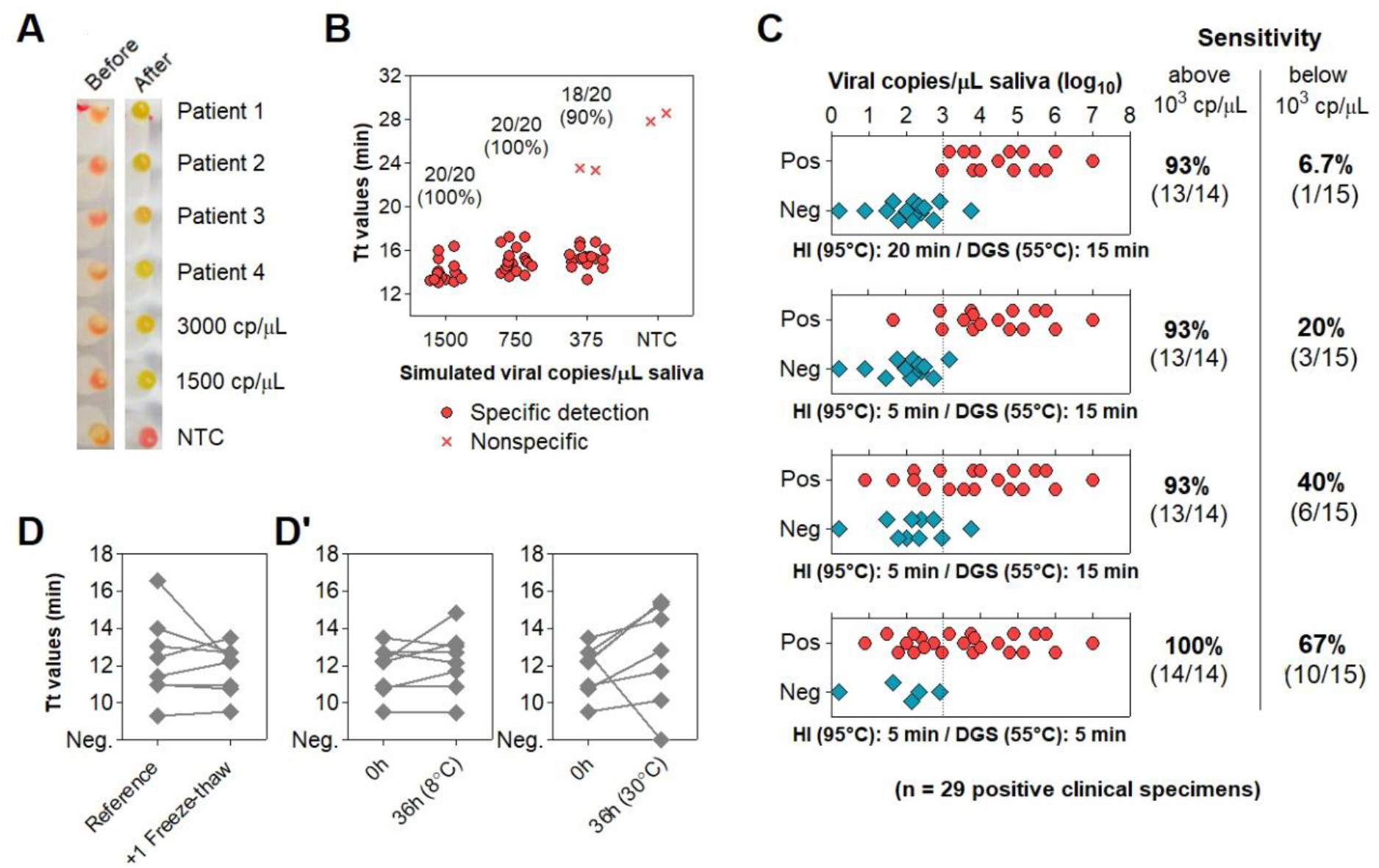
Direct rtRT-LAMP output after volumetric adjustments and reduction of processing time. **A, B)** Output after volumetric adjustments. **A)** Representative color output before and after rtRT-LAMP performed on 4 clinical specimens and on 3000 and 1500 simulated viral copies/µL. **B)** Sensitivity using simulated specimens (20 replicates each). No-template control (NTC) reactions had H_2_O as input. **C)** Reduction of heat inactivation (HI) and DGS incubation times with respective rtRT-LAMP readout and sensitivity (stratified at 10^3^ copies/μL; dashed line). **D, D’)** Before-after plots for 8 specimens processed with HI for 5 minutes and DGS for 5 minutes. **D)** rtRT-LAMP Tt profiles after 1 freeze-thaw cycle. **D’)** Tt profiles after storage for 36 hours at 8°C and 30°C. cp/μL = viral copies per μL of saliva.

To accelerate sample processing, we tested clinical saliva samples confirmed positive via RT-PCR, as better detailed in the next item. We simultaneously processed 29 positive specimens with the previously devised protocol (heat inactivation for 20 minutes at 95°C and DGS stabilization for 15 minutes at 55°C) and under shorter processing times (5 minutes at 95°C and 5 minutes at 55°C). Here, primer set N1 was replaced by E1, which is reported as more sensitive (12) (Table S1). Shortening both heat inactivation and DGS stabilization to 5 minutes led to the largest improvement in detection rate, enabling detection in 100% (14/14) of samples containing over 10^3^ viral copies/μL and 67% (10/15) of samples below that cutoff, a 10-fold improvement over the unmodified protocol (Fig. 4C). These results show that reducing sample processing time improves direct RT-LAMP sensitivity, particularly for low viral loads in saliva.

To assess conservation of diagnostic properties, 8 of these DGS:saliva mixtures were stored at -80°C, thawed and analyzed via direct rtRT-LAMP, without consistent increments in Tt values. Considering that the processed specimens were freeze-thawed twice before this test, this suggests that DGS-processed saliva withstand repeated freeze-thaw cycles (Fig. 4D). Following this, DGS:saliva mixtures were incubated at 8°C and 30°C for 36 hours. At 8°C, no consistent Tt increments were observed, while at 30°C, all samples showed increased Tt values, and one returned negative, suggesting that refrigeration is necessary to maintain diagnostic properties of specimens after processing (Fig. 4D’).

### Characterization of the diagnostic properties of saliva compared to NOP swabs

To evaluate the diagnostic performance of saliva, we analyzed 51 saliva samples collected concomitantly with NOP swabs from symptomatic individuals upon hospital admission. These cases were confirmed by an external laboratory via NOP RT-PCR targeting genes N and RdRp, returning Ct values ranging from 36 to 22.8 (mean_N_ = 29.8, SD_N_ = 3.5; mean_RdRp_ = 30.4, SD_RdRp_ = 3.6). We quantified viral copies in the paired salivas via RT-PCR for the N gene, wherein 48 specimens returned positive (94% agreement). These samples showed viral loads between 1.6 and 10^7^ copies/µL of saliva (0.2-7 log_10_ copies/µL; mean = 3.51, SD = 1.51) (Fig. 5A; Table S2). No correlation between NOP swab Ct values and viral copies in saliva was observed (Fig. 5B). On average, Ct values in saliva were lower compared to NOP swabs (one-way ANOVA with Dunnet post-tests; p<0.05), indicating higher viral load in saliva (Fig. 5C).

**Figure 5:**
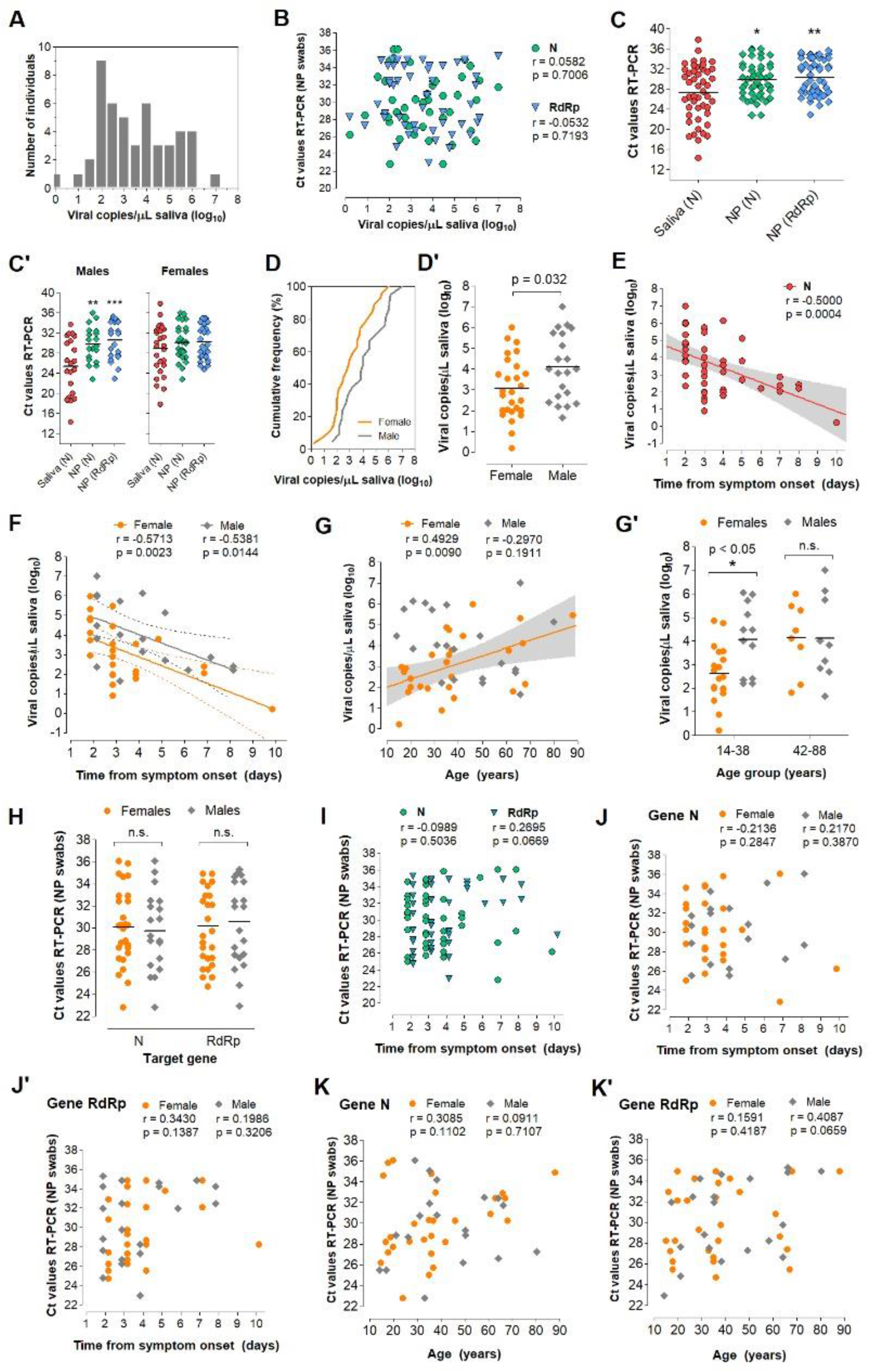
Analysis of saliva as a diagnostic specimen for SARS-CoV-2 detection. **A)** Distribution of viral load in saliva from 51 COVID-19 patients. **B)** Pearson’s correlation between viral load in saliva (gene N) and Ct values of NOP swab specimens (genes N and RdRp). **C)** Comparison of mean Ct values between saliva and NOP swab specimens; and **C’)** the same analysis was performed for each gender. One-way ANOVA with Dunnet’s post-tests; *p<0.05; **p<0.001. **D)** Cumulative frequency distribution and **D’)** comparison of mean viral loads between males and females; Student’s t-test. **E)** Pearson’s correlation between salivary viral load and days since onset of symptoms and **F)** the same analysis stratified by gender. Dashed lines or gray shading indicate 95% confidence intervals of the linear trends for significant correlations. **G)** Pearson’s correlation between viral load and age for each gender. **G’)** Comparison of mean viral loads between genders for younger (aged 14-38) and older (aged 42-88) individuals; Two-way ANOVA with Bonferroni post-tests. **H-K’)** Comparisons performed for saliva **(D’, E, F, G)** were applied to NOP swabs using Ct values from RT-PCR targeting N and RdRp **(H, I, J/J’, K/K’)**.

Next, we analyzed viral load stratified by gender, time from onset of symptoms, and age (Table S2). These clinical data were available for 49 of the 51 individuals. Interestingly, males showed lower mean Ct values in saliva compared to NOP swabs, which was not observed for females (Fig. 5C’). Furthermore, males showed significantly higher viral loads in saliva than females (mean difference = 1.03 log_10_ copies/µL; p = 0.032, Student’s t-test) (Fig. 5D, D’), while no gender differences were found for NOP swab Ct values. In both male and female saliva, viral load peaked during the initial 2 days of symptoms and was negatively correlated with time (range = 2 to 10 days from onset of symptoms) (Fig. 5E). Linear regression analysis showed that salivary viral load falls below 10^3^ copies/µL at around 5 days after symptom onset (x = 4.92 days [95%CI: 3.92 - 6.86]), which in females occurs earlier than in males (x_females_ = 3.74 days [95%CI: 2.42 - 5.24]; x_males_ = 6.35 days [95%CI: 4.71 - 15.38]) (Fig. 5F). Further, viral load in saliva was positively correlated with age in females, but not in males (range = 14-88 years) (Fig. 5G). Notably, among younger patients (aged 14-38), females showed lower viral load than males, while no differences were observed in older individuals (aged 42-88), suggesting that the observed gender differences in salivary viral load decrease with age (Fig 5G’). In NOP swabs, no relationship was detected between viral load (Ct values) and age, gender or days since symptom onset (Fig. 5H-K). Together, these results show that saliva is a reliable diagnostic specimen in comparison to NOP swabs. Moreover, they show that viral load in saliva peaks in the early days of symptoms and is depleted with time, with young females displaying lower viral load compared to males.

### Direct RT-LAMP in 80 saliva samples from symptomatic individuals

Without prior knowledge of NOP RT-PCR results, we tested 80 additional saliva samples from symptomatic individuals collected concomitantly with NOP swabs. Specimens were processed with the established DGS protocol. Direct rtRT-LAMP was carried out with primer sets N1, E1 and As1e, which was also reported to be more sensitive than N1 (Table S1) (12), in addition to the human-specific primer set *ACTB*. All specimens showed successful amplification for *ACTB*, confirming adequate sample quality (data not shown). Reactions using E1 were performed in duplicates and results were considered positive upon detection in at least one of them, while single reactions were carried out with N1 and As1e. Nonspecific and failed amplifications were considered negative. RT-PCR in NOP specimens reported 51 positives (NOP^+^) and 29 negatives (NOP^-^).

Among the 51 NOP^+^ individuals, saliva rtRT-LAMP returned positive in 40 with primer set E1, 35 with primer set As1e, and 31 with primer set N1, which was excluded from subsequent analyses due to reduced sensitivity and excessive nonspecific amplifications (Fig. 6A). Joint analysis considering either E1 or As1e resulted in 41 positives and 10 negatives. RT-PCR on saliva from these 10 discordant NOP^+^ individuals revealed either viral loads below 10^2^ copies/µL or absence of SARS-CoV-2 (Fig. 6B).

**Figure 6:**
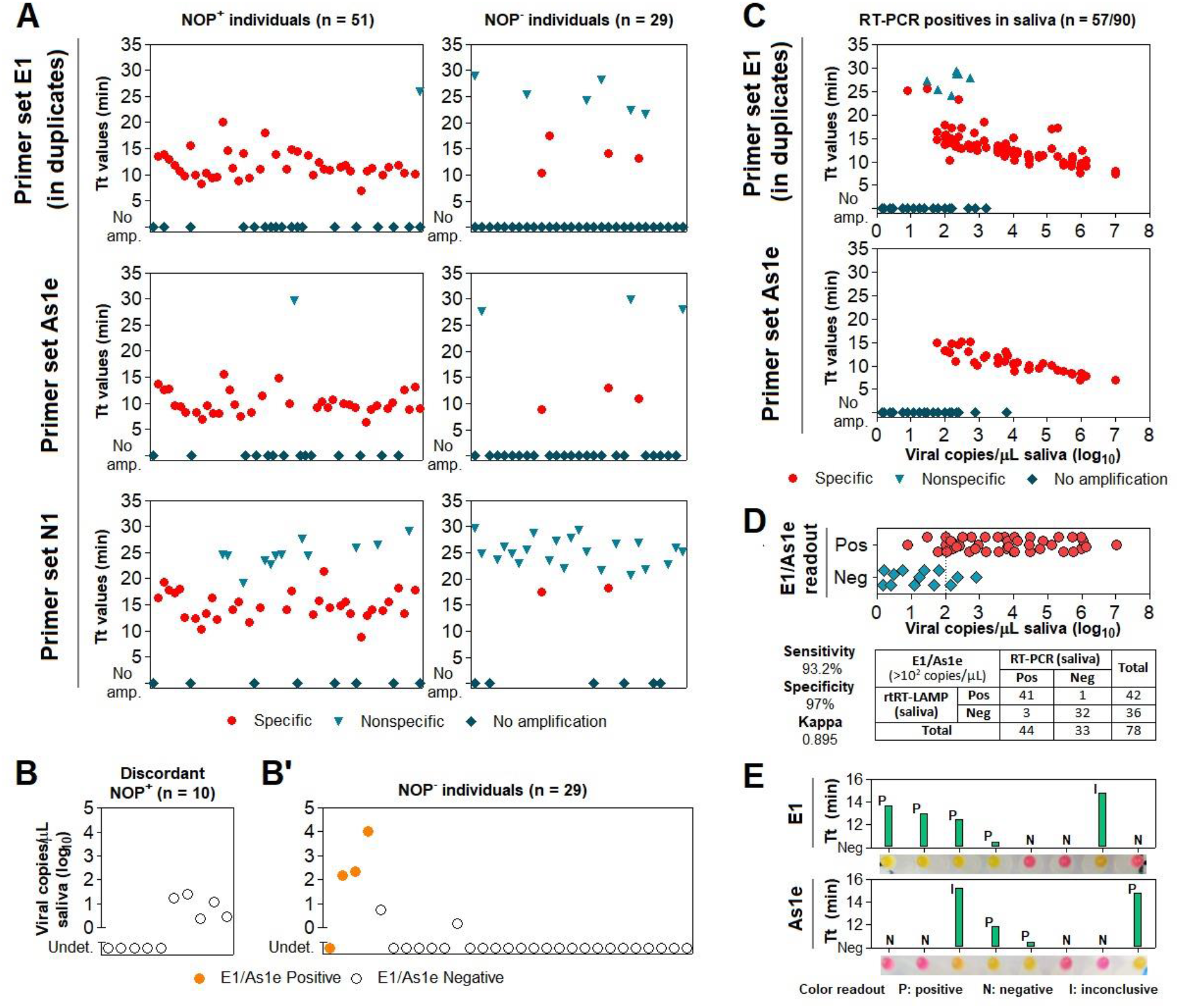
Assessment of the DGS and direct RT-LAMP diagnostic workflow in clinical saliva. **A)** Saliva direct rtRT-LAMP output in NOP^+^ (n = 51) and NOP^-^ (n = 29) individuals with primer sets E1 (in duplicates), As1e and N1. Specific amplifications were classified as positive detections, while nonspecific and failed amplifications were classified as negative. RT-PCR quantification of viral copies in saliva from the 10 discordant NOP^+^ individuals who escaped detection via rtRT-LAMP and **B’)** in saliva from the 29 NOP^-^ individuals, including the 4 discordant rtRT-LAMP positives. Undet. = undetermined. **C)** rtRT-LAMP with primer sets E1 (in duplicates) and As1e on salivas in which viral titer was determined via RT-PCR (shown in B/B’ and in Fig. 5). **D)** Assessment of sensitivity, specificity and Kappa values of rtRT-LAMP considering primer set E1 alone or E1/As1e for specimens containing at least 10^2^ viral copies/µL (dashed line). **E)** Representative comparisons between visual color interpretation and rtRT-LAMP results (Tt values). Over 97.6% of specimens were correctly classified as positive (P, yellow) or negative (N, pink) for E1 and As1e, and the remaining was classified as inconclusive (I, orange-shaded).

Among the 29 NOP^-^ individuals, joint E1/As1e analysis returned 4 positive salivas, which were confirmed in an independent rtRT-LAMP assay (data not shown). RT-PCR on saliva from these NOP^-^ individuals detected SARS-CoV-2 at >10^2^ copies/µL in 3 of the 4 E1/As1e positives. The remaining patients, negative in both NOP RT-PCR and saliva rtRT-LAMP, showed either undetectable (23/25) or low salivary viral load (<10^1^ copies/µL; 2/25) (Fig. 6B’). These results further highlight viral load disparities between NOP and saliva specimens, and indicate that a fraction of infected patients may escape detection via NOP RT-PCR.

### Clinical performance of the direct RT-LAMP workflow

To evaluate the diagnostic capabilities of our workflow, we tested a total of 90 salivas in which viral load was herein determined via RT-PCR, comprising 58 specimens confirmed positive and 33 specimens confirmed negative for SARS-CoV-2. These specimens had been processed with DGS and stored at -80°C. rtRT-LAMP reactions were carried out using E1 in duplicates and As1e in single reactions, as previously described. One positive sample failed to amplify *ACTB* likely due to low quality, and was excluded from the analysis (data not shown). rtRT-LAMP correctly identified 38/57 positive specimens with As1e and 44/57 in at least one of the E1 replicates, with the majority of specimens that escaped detection displaying viral loads below 10^2^ copies/µL (Fig. 6C). Notably, joint E1/As1e analysis also returned 44/57 positives since all 38 As1e-positives were covered by E1, suggesting little benefit from pairing E1 and As1e as presented. Considering the full range of viral loads represented here, this results in 77.2% overall sensitivity and 97% specificity (32/33 negatives) with primer set E1 or joint E1/As1e. Importantly, stratification at 10^2^ copies/µL resulted in 93.2% sensitivity (41/44 positives) and Kappa = 0.895 for viral loads above this cutoff, demonstrating high sensitivity, specificity and reliability of the test (Fig. 6D).

Finally, we sought to determine the efficiency of the RT-LAMP color readout upon visual examination. Without prior knowledge of the results, the color output of 131 samples tested herein via rtRT-LAMP was visually classified as positive (yellow), negative (pink) or inconclusive (orange-shaded), for primer sets E1 and As1e. All yellow- and pink-colored reactions were correctly classified as positive and negative, respectively, resulting in 97.6% agreement for primer set E1 (206/211 reactions) and 97.7% agreement for primer set As1e (128/131 reactions). The orange-shaded output of the remaining reactions resulted from nonspecific or specific amplifications mostly with late Tt values in rtRT-LAMP, indicating absence of SARS-CoV-2 or low viral loads, respectively (Fig. 6E). This demonstrates high agreement between endpoint color output and rtRT-LAMP analysis.

## Discussion

In this work, we developed a DTT/GuHCl-based RNA stabilization solution (DGS) and a workflow that enables robust detection of SARS-CoV-2 in saliva via direct RT-LAMP, with either colorimetric or real-time fluorescence readout. We showed that heat treatment of saliva mixed with DGS protects SARS-CoV-2 RNA from degradation, while improving efficiency, reaction speed and detection rates during subsequent analysis by RT-LAMP. We also characterized saliva and NOP swab specimens according to viral load, gender, age, and time from symptom onset, providing more insight into the advantages and limitations of salivary SARS-CoV-2 diagnostics.

In simulated saliva, we observed that heat treatment with PK-based formulations is insufficient to stabilize viral RNA, contrasting previous reports using samples containing SARS-CoV-2 virions (7,13–15). This discrepancy could be explained by the fact that the spiked RNA in our simulated samples is not protected by the nucleocapsid and other structural viral proteins, being readily digested by any active nucleases left after sample processing. This indicates that PK did not sufficiently inactivate salivary nucleases under the conditions examined here. Thus, care must be taken when using PK and other agents to process specimens, especially when combined with additional viral lysis methods, which may expose viral RNA to digestion. Accordingly, although SARS-CoV-2 remains stable in crude saliva (16), diagnostic sensitivity may drop depending on the method employed to inactivate/process samples, such as heat inactivation, inclusion of detergents, and other factors (17,18), making the RNA protection provided by DGS nevertheless critical.

In clinical saliva, DGS heat treatment improves detection of SARS-CoV-2 compared to heating without DGS. The active ingredients in DGS act by reducing and denaturing salivary extracellular ribonucleases and other inhibitors, and may also facilitate access of RT-LAMP enzymes to the SARS-CoV-2 genome through reduction and denaturation of viral proteins. GuHCl was recently reported to improve speed and sensitivity when added to RT-LAMP (10), so carryover of DGS into reactions may cooperate to increase SARS-CoV-2 detection by modulating reaction chemistry as well. Importantly, this carryover is not solely responsible for the rise in detection rates and reaction speed in clinical specimens, because these effects were observed in relation to no-DGS controls in which RT-LAMP was supplemented with GuHCl (40 mM). Thus, DGS improves SARS-CoV-2 detection both via stabilization of the viral genome and increasing RT-LAMP speed/sensitivity. Furthermore, DTT/GuHCl are also mucolytic agents (19), and therefore ameliorate pipetting of viscous specimens.

Of the DGS protocols investigated here, the best overall performance was achieved with a 2-step method that includes heat inactivation of the saliva in the collection vessel before further manipulation (protocol B). This protocol improved direct RT-LAMP speed and detection rates compared to the no-DGS control, and elicited the strongest RNA stabilization effects compared to the other methods we examined. This 2-step protocol reduces risk of infection by healthcare specialists and clinicians since the collection vessel remains sealed until heat inactivation of the virus. Therefore, it facilitates diagnostic workflows and alleviates biosafety concerns in point-of-care settings and in test sites lacking in sophisticated infrastructure.

Additional modifications to protocol B improved the diagnostic capabilities of the test. Color discrimination can be challenging due to interindividual variations in specimen quality, such as salivary pH. The volumetric adjustments to the DGS:saliva ratio and RT-LAMP mastermix improved endpoint color interpretation, while shortening the heat inactivation and DGS steps to 5 minutes each improved sensitivity for specimens containing less than 10^3^ copies/µL, likely owing to greater RNA stability. These steps should suffice to inactivate SARS-CoV-2, since complete viral inactivation has been reported for periods as short as 3 minutes at 95°C (20,21). Furthermore, the final sample processing protocol is amenable to repeated freeze-thaw cycles and allows storage at 8°C for up to 36 hours without adversely affecting diagnostic output, which is important if RT-LAMP must be performed or repeated later. Finally, shortening sample processing time (from 35 to 10 minutes) further reduced diagnostic turnaround time.

After these modifications, RT-LAMP achieved 77.2% overall clinical sensitivity and 97% specificity, with 93.2% sensitivity in saliva containing at least 10^2^ viral copies/µL (Kappa = 0.895), which is on par or more sensitive compared to most of the direct RT-LAMP approaches reported so far (7,12,22–30). Moreover, the high agreement (>97.6%) observed between color interpretation and specific amplification plots in rtRT-LAMP demonstrates reliability on end-point color readout if real-time analysis is not possible. Although only 23% of salivas containing less than 10^2^ viral copies/µL were detected by the present method, the gain in exam turnaround time in the present method may be desirable at the cost of sensitivity, since lower viral loads are associated with lower transmissibility of SARS-CoV-2 (31). Likewise, it has been proposed that effective COVID-19 surveillance depends on test frequency and turnaround time rather than on test sensitivity (32), especially for identifying nonsymptomatic carriers. Therefore, periodic testing using a faster, cheaper and noninvasive saliva protocol may be preferable to more lengthy and costly methods, such as standard RT-PCR workflows.

Using RT-PCR, we detected SARS-CoV-2 in 94% of the salivas from symptomatic patients, with Ct comparisons suggesting that viral load in saliva may be higher than in NOP specimens, particularly in males. The observed absence of correlation between NOP and salivary viral load reflects distinct viral shedding dynamics in these tissues (2). Our results confirm that saliva is a suitable diagnostic specimen compared to NOP swabs, especially in cases where SARS-CoV-2 is present only in saliva, as ascertained in 3 patients herein. Male sex and old age are risk factors for developing severe COVID-19 (33,34). Here, males showed mean viral load around 10 times higher than females in saliva but not in NOP swabs. Since airborne transmission is related to the amount of virions per saliva droplet, this indicates that males could be more likely to spread the virus than females. Notably, compared to age-matched males, young females (< 38 years old) showed lower salivary viral load that increases with age, while no clear age-related effects were found in males. Clinical severity and immunological profiles seem to better correlate with viral load in saliva than in NOP swabs (35,36), so our findings could be attributed to distinct immune responses between genders leading to higher viral shedding and disease severity in males (37), and could also explain the higher proportion of asymptomatic females in couples positive for SARS-CoV-2 (38). These observations further suggest that salivary viral load together with older age, male gender and other risk factors could be important to predict disease duration, severity and mortality (36,39). Viral load in saliva peaked in the first days of symptoms and declined with time, agreeing with recent estimates that show peak transmissibility 1.8 days before onset of symptoms and low chance of transmission beyond 9.5 days after onset (40). Importantly, we estimate that the sensitivity threshold of 10^3^ copies/µL employed by several RT-LAMP approaches is crossed around the 5^th^ day of symptoms, with males showing delayed virus clearance compared to females. Based on these observations, we suggest that salivary diagnostics in symptomatic individuals should be performed as soon as possible after symptom onset to increase chances of detection, especially for young females. Recent reports show that saliva has higher sensitivity than nasal/nasopharyngeal swabs to identify asymptomatic cases (41), and that viral load distribution is equivalent in saliva from symptomatic and nonsymptomatic individuals (42). Therefore, rapid salivary diagnostics stands as an invaluable opportunity to efficiently monitor and curb SARS-CoV-2 transmission.

In summary, we report a simple and rapid RT-LAMP diagnostic workflow that obviates the necessity of RNA extraction and specialized equipment, providing a cost- and time-efficient alternative to standard RT-PCR diagnostics. The use of DGS to process specimens and modifications to the RT-LAMP reaction resulted in high sensitivity and specificity to detect SARS-CoV-2 in saliva via real-time or endpoint colorimetric analysis, thus qualifying it for point-of-care testing. Our results not only confirm that saliva is a good diagnostic fluid, but also indicate that it may provide clues to clarify the biological determinants of SARS-CoV-2 infection and transmission. Finally, we reinforce that salivary diagnostics should be prioritized in screenings to identify presymptomatic and symptomatic cases of COVID-19, which may be extended to other respiratory pathogens.

## Data Availability

The data that support the findings of this study are available from the corresponding author, MRPB, upon reasonable request.

## Acknowledgements

We are grateful to all patients who participated in this study, and we hope that you and yours can quickly recover from this challenging situation. We also thank all healthcare professionals, researchers and others combating the COVID-19 pandemic in Brazil. We thank Prof. Edison Durigon (ICB/USP) for providing inactivated SARS-CoV-2 virions and the staff at HUG-CELL and Instituto de Biociências/USP for enabling and supporting this work. Funding was provided by FAPESP (20/05949-2), FAPESP/CEPID (2013/08028-1), JBS, and Itaú-Saúde para Todos.

